# The potential role of cultural and religious healing practices in shaping community vulnerability to highly infectious diseases in western Kenya

**DOI:** 10.1101/2024.04.24.24306297

**Authors:** Naomi Nga’ng’a, Reuben Onkoba Momanyi, Caleb Chemirmir, Hazael Biwott, George Ayodo, Monica Orero, Damaris Ochanda, Sarah Ngere, Winnie Ogola, Tutus Murundu, Geoffrey Munene, Zachary Misiani, Michael Ayaibei, Richard Dimba Kiaka

**Affiliations:** Kenya Red Cross Society; Jaramogi Oginga Odinga University of Science and Technology (JOOUST); Kenya Medical Research Institute (KEMRI); Masinde Muliro University of Science and Technology (MMUST)

**Keywords:** Traditional healing, religious healing, looping effect, community vulnerability, highly infectious diseases, western Kenya

## Abstract

We draw from empirical research conducted in communities in three border counties in western Kenya – Homa Bay, Bungoma and West Pokot - to examine how cultural and religious beliefs and healing practices can potentially shape the vulnerability of those communities to highly infectious diseases. Fieldwork consisting of mixed methods namely, key informant interviews with traditional and religious healers as well as their patients, focus group discussions with community members knowledgeable on cultural customs and practices, and participatory inquiry workshops with health professionals and administrators were used to collect the data.

We find that traditional and religious beliefs and healing practices potentially shape community vulnerability to highly infectious diseases in two major ways. First, is a dualistic illness etiology involving a biomedical and socio-cultural etiology. Unexplained illnesses and illnesses that did not respond to conventional medicine were treated using traditional medicine. Making traditional and religious healers the first mile treatment preference could potentially delay appropriate treatment and compromise safe handling in case the disease is highly infectious. Second aspect pertains to the risks in the traditional and spiritual healing practices. Shared consecrated water often fetched from a large water body, laying of hands, use of herbs and rituals involving slaughtering of animals enhanced contacts. The use of protective gears during healing was inconsistent among the healers, but largely lacking as many healers could not afford them or were considered to reduce patient’s faith in the healing powers. These practices potentially predispose people to highly infectious diseases and can hasten the spread and symptom severity. To reduce the vulnerability of border communities to highly infectious diseases, we argue for a need for comprehensive strategies that consider the intersecting factors of vulnerability to outbreaks, healing beliefs and practices. This may involve policy initiatives aimed at integrating traditional medicine practice and the mainstream health system.

## Introduction

Cultural and religious practices and beliefs around the world have significant influence on the management of outbreaks of infectious diseases, where swift response and effective control of the diseases is required (McMichael 2004; Manderson 1998; Borg 2014). While some cultural and religious beliefs and practices can complement effective response to and management of highly infectious diseases (Rodriguez-Dod et. al. 2016; Victor and Ahmed 2019), others inspire health seeking behaviors that can be counterproductive to interventions. This has been exacerbated especially in situations where the well-intended interventions by emergency organizations and governments are in disconnect with communities’ cultural practices of seeking health and responding to epidemics. Recent examples such as response to COVID-19 (Alwala 2020; Oey and Rahardjo 2021; Okafor 2022; Lucy et. al. 2022) and Ebola Virus Disease (EVD) (Shultz et al. 2016; Park 2020) offer relevant testimonies to the conflicts between communities and government’s interventions to manage the infectious diseases. The containment of these highly contagious diseases require stringent measures that interfere with the daily activities of the members of the communities including, for example, restricting social interactions and movements of people, and suspending important cultural ceremonies and rituals (Alwala 2020). Through policy actions such as movement restrictions (or lock-down), government safe burial procedures amongst others, governments and other health emergency organizations take control of aspects of people’s lives that is normally a preserve of cultural and religious doctrines. Communities can often violate or resist these intervention measures that tend to challenge the role of culture and religions in people’s lives. Whether the resistance is active or passive, it constrains government’s interventions, thereby compromising health outcomes.

Blaming and victimizing cultures and religions of communities for the contradictions that emerge in these emergency health interventions is common (Jones 2011; Wona 2018). Governments often see the traditions, especially those they consider to be unique, to be an impediment to health interventions during outbreaks of highly infectious diseases (Kipiriri and Ross 2020). These policy attitudes only help to recast the conflicts between formal interventions with cultural practices. Medical anthropologists have asserted that it is imperative to consider very actively the way local cultures interact with the concept of health, if we are to make any significant reconciliation between our formal health interventions and cultural practices around health (Airhihenbuwa et al. 2014; Manson 2020). That is, before asking a community to assume particular health habits, it is necessary to ascertain the existing health habits of the people and the interlinkages between those habits, their functions and the meanings they inspire amongst the people practicing them (Chiavo 2013). Such an approach will require a more radical paradigm shift from top-down health interventions to a more community centered-approach, where local health behaviors and how they are rationalized by the community members are at the core of analysis. Importantly, studies should consider understanding how the cultural practices shape and are shaped by formal health interventions, especially in the context of highly infectious diseases when socio-economic aspects of the communities are often under pressure and health resources are overstretched.

Many communities across Africa use non-biomedical treatments to promote health, including herbal medicine preparations and spiritual healing (Harrison and Airhihenbuwa 2018; Adu and Anderson 2019). Reasons could range from lack of adequate access to health facilities, high costs of biomedical treatments and traditional and religious beliefs around diseases and health. Even with the rising promotion of western medicine, the use of traditional and religious healing has remained an important source of health for many communities to the extent that the traditional and religious healers are the first stop for many people in health-seeking for highly infectious diseases like EVD (de-Vries et al. 2016). Experience from Africa has shown that when disease intervention programs fail to recognize and work with indigenous beliefs and practices, they also remain largely ineffective and miss their goals (Geissler 2005; Boahen 2013). The World Health Organization (WHO) in its *Traditional Medicine Strategy 2014 – 2023* has emphasized the global importance of these alternative approaches to health (WHO 2103). In recent decades, research in medical anthropology, sociology and other social health sciences have promoted broader approaches that accommodate non-western health-seeking. That is, they embrace a shift from rejecting cultural and traditional health beliefs and practices to understanding that culturally rooted, traditional healing approaches are valued and used by many people often in combination with biomedical approaches. In order to understand why infectious diseases programs are not working and endeavor to provide solutions, research and interventions must consider the culturally embedded health seeking behaviors.

Drawing on fieldwork conducted in selected communities in western Kenya, we undertake an analysis of how some traditional and religious beliefs intersect with the concepts of disease, health and healing. First, we examine the factors that shape health related-beliefs and practices, including seeking healing from traditional and religious healers. We then analyze how the practices can influence management of highly infectious disease including community preparedness, response, surveillance and control of highly infectious diseases. We borrow the concept of looping effect to argue that through cultural or religious beliefs and practices, communities form loops with the bio-physical disease, so as to redefine meanings of treatment and overall goal of good health. Considering that highly infectious diseases spread very fast and therefore require swift interventions, grounds the value of evaluating those loops to broaden and sharpen an understanding of the barriers and enablers of cultural knowledge and behaviors in preparedness, planning and execution of emergency response in times of outbreaks of highly infectious diseases, which still falls short of the mark in many epidemics management programs.

We proceed to describe our theoretical framework where we situate our work in the literature in medical and cultural anthropology, as well as cultural psychology that have applied the concept of looping effect in understanding how culture shapes health behaviours and associated diseases epidemiology. Next, we describe the methodology including a description of the study sites and data collection and analysis methods. We then present our findings and discuss them in light of their implications to scholarship, policies and the practices for intervening on highly infectious diseases outbreaks. We finally draw some conclusions and suggest some recommendations.

### Analytical framework/theoretical framework

Our contribution in this paper pays close attention to the social and behavioral aspects of infectious diseases within communities where non-western cultures and religions strongly mediate social life and understanding of health. We therefore tap from the theoretical inspirations from medical anthropology and cultural psychology to attend to the questions of how culture, mind and biology interrelate within the domain of infectious diseases management (Schaller and Murray 2010). Community experience and response to highly infectious diseases can be enmeshed in the sociocultural beliefs and practices that mediate their interaction with the physical and metaphysical world (Schaller and Muray 2010). In particular, the social transmission of highly infectious diseases means that their spread, and hence their impact on a community, is driven in part by social behaviors which, in turn, are shaped by patterns of culturally shared beliefs in that community (Baye et al. 2021). We particularly draw from the conceptual insights of *looping effect,* originally coined by Hacking (1995, 2007), to analyze how the interrelations between cultural beliefs or practices and psychological processes can inspire certain behaviors and responses towards highly infectious diseases in ways that have implications on disease control and the disease itself (Kirmayer and Sartorius, 2007; Ryder and Chentsova-Dutton, 2015). That people in a community can choose to employ traditional and religious healing in response to highly infectious diseases does not hinge on a conceptual or cognitive vacuum. Rather, their choice for health-seeking behaviours are a result of various kinds of socially embedded knowledge in the unique cultural and historical settings they inhabit. Hacking calls this process ‘dynamic nominalism’ through which cultural institutions, social contexts, and socially embedded knowledge produce various kinds of looping effects to the biology of diseases (Hacking 2007). With this framework, we connect how religious and traditional healing becomes a preference in communities we studied, and how the health-seeking behaviour can form looping effects with highly infectious diseases and responses to them. Therefore, instead of seeing culture – the metaphysics, and disease-causing pathogens – the physical as two parallel existences, the looping effect allows us to analyze the indirect mutual interaction between the two worlds (Baye et al. 2021). Looping effect considers that while people’s beliefs “do not intervene directly on the virus itself and are not so clearly implicated in the core symptoms of the infection” they can change important features of the highly infectious diseases such as infection rate, severity of infection and tolerance and resistance to drugs through virus mutations” (Baye et al. 2021:3).

In advancing this thought, we consider that three cultural and psychological aspects of highly infectious diseases can create a looping effect with the biology of the diseases. These three aspects include diseases etiology, treatment responses and perceptions on institutional responses. Etiology is concerned with how people conjure diseases, including how they give meanings to their existence or causality. What people believe to be the cause of a disease will determine their response to the disease (Balde 2016). Many African communities are endowed with cultural repositories of knowledge and experiences from which they draw meanings of diseases and their causes. These can range from metaphysical, relational and environmental causes. Individuals in the community will seek-health for diseases in a manner commensurate to causes of those diseases. Secondly, health-seeking behaviours can inspire diverse implications on societal responses to an outbreak, including delaying appropriate treatment, weakening surveillance and reporting, enhancing spread and severity. These can further make the pathogen to be resistant to treatments. We use the concept of looping to address two questions. 1). How can traditional and religious beliefs and associated practices form a looping effect with highly infectious diseases and their treatments? 2). What implications can they have on the effective management of the disease and ensuring good health in case of disease outbreak?

## Methodology

### Study settings

Fieldwork conducted in western Kenya in the counties of Homa Bay, Bungoma, and West Pokot. In the context of highly infectious diseases, the three counties share a porous border with neighboring Uganda and Tanzania which have in recent times seen outbreaks of Ebola Virus Disease (Kinsman 2012; Kozlov 2022) and Malburg (Deb et al. 2023) respectively. Homa Bay County has also struggled with multiple outbreaks of cholera in recent years.^1^ Moreover, these counties are characterized by strong traditional belief systems that influence health seeking behaviour (Chebii et al., 2020; Kwanya, 2020; Mbuithia et al., 2018; Otieno and Onyango, 2018). There is also a persistent practice of unique religious beliefs including deifying humans and asserting that healing can possibly be achieved through faith and religious rituals e.g. in Bungoma. These religious beliefs influence health seeking for communities in ways that impinge on management of highly infectious diseases, including EVD. Lastly and equally important, these counties were selected for surveillance and health education by a national taskforce on EBVD following the 2022 outbreak in Uganda.

## Methods

Fieldwork was conducted between September and November 2023. Qualitative data was collected using a combination of methods including key informant interviews (KIIs), focus group discussions (FGDs) and participatory inquiry workshops. KIIs guides were developed and administered a constitution of a less structured, open-ended set of questions organized in pre-identified thematic areas. A total of 45 were conducted including traditional healers (13) and religious healers (16) and their respective patients (16) in each of the counties. Participants were identified through chain referrals. Interviews were conducted within health facilities or respondents’ homes depending on the comfort of the respondent. Interviews were conducted in a language that the respondent was most comfortable with. Whereas in West Pokot county the interviews were translated, in Homa Bay and Bungoma, the researchers were able to directly conduct interviews in Dholuo and Swahili respectively.

FGD guides were semi-structured providing respondents with the flexibility to express their opinions. A total of 6 FGDs each (8-12 participants) were conducted with community members in the three counties. Each FGD was of a homogenous gender in that women FGD was separated from Men FGD. Respondents were brought to a central point where sessions were conducted. The FGDs investigated a set of issues on sociocultural and religious healthcare seeking on highly infectious diseases. In Homabay Dholuo was used to facilitate the discussions, in Bungoma Swahili was used to facilitate the discussions and lastly in West Pokot a translator assisted the moderator to translate questions into Pokot.

Participatory inquiry workshops were used to collect data and information from stakeholders of public health promotion stakeholders in each county. These stakeholders included government public health officers, NGOs and CBOs, Community Health Promoters (CHPs) and national administration officers – chiefs. The workshop was conducted in central locations. Participants were divided into groups depending on their category. Each group had a moderator and a note taker. They were given a list of questions to discuss and generate detailed group responses. After about an hour of group discussions, participants gathered in a plenary to discuss their points. Points were interrogated by participants and adjustments made as was relevant and agreeable to participants.

All discussions from KIIs, FGDs and participatory inquiry workshops were audio recorded and transcribed verbatim into English. The framework method was employed to analyze the data, using both inductive and deductive coding. The authors jointly developed a coding framework that was jointly developed and formulated into a spreadsheet matrix that contained numerous themes and sub-themes under which relevant excerpts from the transcripts were entered. Irrelevant codes were deleted while adding emergent codes to the matrix as needed. Transcripts were read again and again to ensure that coding was done exhaustively and also checked for consistency. Emerging themes were collated, collapsed, defined and interpreted as results.

### Ethics Statement

Ethical approval No. MMUST/IERC/185/2023 was granted by the Masinde Muliro University Institutional Scientific Ethics Review Committee (MMUST-ISERC). Respondents’ informed consent were sought in writing before interviews. Under the informed consents the respondents were provided with all the pertinent information about the study to guide them in making a decision on whether to participate in the study or not. The information provided in the consent forms included: the purpose of the study, the procedures to be followed; and the benefits/risks of this study. There was no foreseen harm to the participants however, participation in the study was completely voluntary, and respondents’ confidentiality has been maintained such that no information in the processed data and publications can lead to the identity of an informant. Data collection took 3 days in each of the 3 counties, where in Homabay the team collected data between 25^th^-27^th^ September while in Bungoma and west Pokot data collection was conducted between 11^th^-13^th^ October 2023.

## Results

### Traditional and religious illness and healing beliefs

#### Beliefs on causes of illness

There exists a dualistic illness etiology in Homa Bay County, Bungoma and West Pokot County. Both traditional and religious (especially those from African Instituted Churches) spheres in the county consider diseases to be associated with biomedical and sociocultural causes. Using a biomedical lens to explain disease etiology, religious and traditional healers, their patients and participants of FGDs associated some diseases to viruses, bacteria and dirt and poor hygiene. For example, cholera was associated with poor sanitation, while the COVID-19 was mentioned to be caused by coronavirus. A respondent elaborates below how to prevent diarrheal diseases

> *These infectious diseases are diarrhea and vomiting. When you hear that there is an outbreak of diarrhea in Luanda that means that it has already spread to these parts of Ang’iya. If you hear about a cholera outbreak in Luanda, people are advised to boil drinking water we fetch from the lake, eat clean food, eat well cooked food, don’t take cold meals and no open defecation, use the toilet. Wash your hands after going to the toilet and dry your hands with tissue paper and throw it in the toilet. When you throw the tissue paper outside and not in the toilet, it helps to spread the disease.*
>
> *Religious Healer KII4, Homa Bay*

Socio-cultural illness etiology is largely associated with customs and beliefs. Witchcraft was mentioned as one of the causes of illness. Witchcraft distinguishing signs are diverse but largely associated with abnormal illness such as mental disorder, bloody vomit, severe headaches and severe stomach aches. Among discussants in a Focus Group one of the respondents says that:

> *…then for example when a child is looked at badly (evil eye) with a witch(iro), then he is taken to the hospital and injected, he will not be well. So some people believe if I run to the religious healer, they will be well*
>
> *FGD2, Homabay*

A respondent explains elaborately how a child can get sick from an ‘evil eye’ in the excerpt below. It is believed that diseases caused by an ‘evil eye’ or witchcraft cannot be treated in the hospital also referred to as *non-hospital diseases*. *Non-hospital* cases are treated by either traditional medicine or by religious healers. The rationale is that if the disease is treated in the hospital the sick person will not recover from the sickness.

> *For example, there is a particular illness that forces you to go for that type of treatment. When someone looks at a child who is breastfeeding with bad eyes; so there are people with bad eyes that will make this milk turn into something else in this child’s stomach. So a hospital won’t help you in this case. So there are people who can scratch the bad milk from inside the child (saro) hence this traditional healers, there are places where they come in handy like such not that they don’t help completely*.
>
> *FGD2, Homa bay*

Disease etiology is also arrived at when a particular disease does not respond as quickly as desired to biomedical treatment. Sometimes when traditional or religious healing is sought, the rationale is driven by non-response to biomedical treatment. Those diseases that take too long to recover are believed to be caused by the power of *satan*. The *satanic* forces are believed to be inflicted by humans in a way of punishing the inflicted person/or when they are envious of the person. In Homa bay a respondent stated that:

> *A sickness that has been treated and has taken even 3 months and it is still that one sickness, it goes and comes back and goes and comes back that now shows that satan is in there so we have to pray to God*.
>
> *Religious Healer KII1, Homa bay*

A patient of a religious healer from Bungoma believes that she was not responding to biomedical treatment because her illness was inflicted by an ‘evil eye’. The respondent believed that an envious person cast a spell of bad luck on them and made them severely ill with a disease that did not respond to conventional medicine.

> *What I can say is “hasad” someone is not happy with your progress and looks at you with an evil eye because they are not happy with your growth. “Hasad” is bad lack.*
>
> *Patient of a religious healer KII2, Bungoma*

In West Pokot a respondent attribute illness causation to supernatural forces that is caused by supernatural forces emanating from the river. When a respondent was asked what causes pneumonia their response was as illustrated below.

> *This disease originates if you cross rivers with supernatural thunders as we were told long time ago. And sure this supernatural being are there because if you cross the river and it doesn’t like you it will throw some soil at you. Last year it attacked some of our children in River chematong where they droned and they were searched for a very long time until one of the person identified them and for sure this supernatural thunders may exist.*
>
> *Traditional Healer KII3, West Pokota*

Some respondents believe that the departure from indigenous traditions such as their traditional food practices is causing the community illnesses. They believe that the food they consume currently makes them susceptible to illnesses.

> *In the past people never used to get sick from these illnesses they used to take this vegetable called mhalaha, it helped their body to be strong, the soup from the cattle heads, they never used to be sick, they used to eat traditional food and now we eat meat from treated cattle, they put chemicals in milk we drink and the diseases are caused by these things*
>
> *FGD1, Bungoma*

Traditional medicines are believed to protect someone from illnesses failure of using these drugs are causing illnesses in people.

> *If you are used to taking traditional medicine, it is very hard for you to contract the diseases because the medicine washes all the blood veins you cannot get infectious disease. It helps you. FGD1, Bungoma*
>
> *Traditional and religious healing beliefs*

Traditional,religious and spiritual beliefs were noted to offer support and meaning to those coping with health challenges. Most of our respondents who went to traditional or/and religious healers were comforted and reassured of healing especially in instances where conventional treatment failed. In religious healing, faith was paramount for healing to take place.

> *What I can say is that I had faith because I knew that when I go there/I did not have a second thought. I did not have that; I knew if I go to the hospital I will get help. But I did not think about what if the Imam comes and pray or read the Quran is when I will get healed, I knew that if I go to the hospital I will get healed. So when I was tired of going to the hospital I told myself to try God may be God will help. So I had faith and asked him to come and pray for me and also I pray for myself maybe through God everything will be fine.*
>
> *Patient of a religious healer KII2, Bungoma*

Depending on the individual one may choose traditional or religious healing.

> *The traditional healers are there hence they treat illnesses that can’t be treated in the hospital, very many illnesses defeat doctors in the hospital hence the traditional doctors are the ones that clear them and once you take your patient there, they begin to drink porridge and they start speaking.*
>
> *Patient of traditional healer KII4, Bungoma*.

In Christianity the doctrines in these churches emphasize the possibility of God giving healing powers to members of the congregation. They receive the healing powers through the holy spirit. Although the doctrines may differ in some areas, these churches share foundational beliefs in some diseases occurring because of curses, sins of witchcraft. Diseases associated with these cultural processes were reported by religious healers to be often associated with strange signs and symptoms that are diverse. One respondent states that God heals because everything happens in his power and therefore healing also comes from him.

> *Everything happens through the power of the holy spirit or God. We believe in prayers and the Religious Healer can pray and we believe through the strength of Jesus Christ the patient is healed.*
>
> *Patient of a religious healer KII2, Homa bay*

The religious healer therefore becomes the vehicle in which healing is performed but God is the healer as stated by a Bungoma respondent

> *We do not say that somebody heals, it is God who heals not an individual. Maybe there is that bit of a praying, there is that bit of reading the verses of the Quran to get that healing from God.*
>
> *Patient of a religious healer KII2, Bungoma*

The Christian religious healers, largely consist of African Instituted Churches e.g., *Roho* and *Legio Maria* churches. These churches combine Christian teachings with Luo customs to advance spirituality. Therefore, it is no wonder that some traditional healers mentioned that some religious healers refer patients to them to collect medicines for the further treatment and cultural advice or send to the hospital for further treatment. Religious healers who were interviewed believe that their powers come from God. It is from these powers that they are able to discern *hospital* and *non-hospital* illness and refer accordingly.

> *God gave me some gift that by just seeing you the way we have sat and when I just start praying for you, I get a vision of what’s going on with you and even in your family. So that’s why I have to do the checking (diagnosis)for me to know the disease was caused in this way and what brought it. And if it happens that the disease was caused by a human, then we will pray to God. But if it’s just a disease which have just come up, then I will send you to the hospital.*
>
> *Religious Healers KII5, Homa Bay*

Religious healers are cognizant of the extent of their ability to treat diseases. Using their judgment and powers given by God traditional healers use a combination of spiritual healing and traditional medicine. In other cases, they refer their patients to the hospital especially for infectious diseases because they appreciate the severity of the disease.

> *The patient was weak and had persistent cough accompanied by pain in the lungs. When he came, I prayed for him and after one week, I sent her to the hospital to check, the tests showed he had no TB.*
>
> *Religious Healer KII3, West Pokot*

### Traditional and religious healing practices

#### Religious healing practices

Religious healing involves rituals that are accompanied by prayer, laying of hands and consecration of the body using *holy water*. According to religious healers and their patients who were interviewed, *holy water* can be sourced through different means. Communities living near large water bodies such as the ones from Homa bay, water bodies and their water are significant in healing practices. the practice include fetching water from Lake Victoria boiling and consecrating it through prayer by a religious leader with healing powers. *Holy water* can also be drawn from water bodies with religious significance or mythical in Luo cultural cosmologies. A religious healer from Homabay explains how she prepares *holy water* in the excerpt below:

> *we boil the water as we pray asking God to bless the water to turn into medicine. So, when I sprinkle in his body you see the body shakes. It sends away the devil. So ill pray for you that way and after that you’ll be healed and taken by your family members*
>
> *Religious healer KII6, Homabay*

In Islam the religious practice involved citation of specific Quran verses, burning incense and praying over the sick person. This ritual is repeated until the sick person recovers.

> *The prayers happened in our home but the first time he burnt incense in his home and my father did the same, he is also a Maalim. It’s just incense, while reading the Quran.* Patient of a religious healer KII2, Bungoma

In West Pokot religious healing involves prayers only, the religious healer lay their hands on the sick person and if they have faith in God then they are healed..

Some processes of administering healing that were mentioned involved body washing, where the healer washes the patient’s body using *holy water* or is washed using a cocktail of herbs mixed with water. The processes involve direct body contact between the healer and the patient.

> *when they come, I will boil that water as I pray vehemently and after that, I first wash them with it after adding salt into it and a bit of sugar to stop the bleeding and can be done three or four times then drinks. He begins to urinate the infections like they urinate blood. At that time, I’ll be praying as I am washing him until the minute he arises.* Religious Healer KII6, Homabay

#### Traditional healing practices

Traditional healers in Homa Bay, Bungoma and West Pokot County are similar and diverse in nature. In the 3 counties they differ depending on their specialty on the kind of diseases they treat. There are those dealing with broken bones and displaced joints, some deal only with mental disorders, others deal with snake bites. There are those who deal with diseases associated with witchcraft, while others deal with diseases that are clinical in nature such as measles, asthma, stomach aches etc. Healing for diseases associated with witchcraft and those that are clinical in nature are the ones that have direct implications on management of highly infectious diseases. A patient of a traditional healer explains how he was treated when he could not find treatment for an illness that was associated to socio-cultural etiology.

The treatment included traditionally cutting through the flesh *(chanja)* and inserting herbal medicine.

> *Because he saw my body swelling, my leg swelling, my chest swelling, he just started making small cuts and inserting medicines and those medicine that got into my body, made me feel pain when they were entering hence from there, it just took me one week and I started seeing my body going back to normal, he started from there then he gave me another one for drinking and I got well after also the herbs which he cut into my body*.
>
> *Patient of traditional healer KII4, Bungoma*

Traditional healers also treat illnesses that can be bio medically defined such as malaria, Typhoid, TB, hepatitis B, sexually transmitted infections and many others. They report that they are able to know what disease the patient has by report from patient, observation of symptoms or palpating the patient. The herbs used are used to specifically treat certain ailments. The treatments are sometimes accompanied by rituals such as slaughtering an animal. In the excerpt from West Pokot below the patient was ‘diagnosed’ with an STI, the treatment included using herbal medication and slaughtering a goat to be part of the treatment process.

> *I gave them ‘Yowoo’ a herb called ‘yowoo’ which she would go and boil for herself at her home. Also I gave a herb called ‘Kelkelowo’ for sexually transmitted diseases. It is those two herbs that she can use but for those who are going to slaughter a goat, they will combine ‘songogoghwo’,’manampellion’ ‘cheporion’. These herbs can be mixed with soup boiled from goat meat. I also instructed her to come back again after some time to examine the blood level because I saw there may be less blood level. She should come back in order to see if the low blood level has retained or it has returned to normal*.
>
> *Traditional healer KII3, West Pokot*

Special ritual is made for protection against diseases in the recent past the communities of West Pokot were cleansed for protection against COVID 19. The ritual involves slaughtering a cow and burning it. Herbal medication is prepared and drunk in order to protect them from the illness.

> *…during cleansing we curse these diseases so they can’t continue they stop there. So, the cleansing is like slaughtering a cow and burn it. Then there are drugs we take, there are others we drink and pour so the diseases don’t come to us in our village.*
>
> *FGD2, West Pokot*

#### The practice of administering healing

Some processes of administering healing that were mentioned to be having the risks of predisposing healers and patients to infection included body washing, where the patient washes the body or is washed using a cocktail of herbs mixed with water. The processes involve direct body contact between the healer and the patient, inhalation, where the patient inhales medicines under a blanket. The blanket is often used by other patients as well; piercing or cutting skin or veins to suck out what was termed as “*bad blood”*, with the high risks of creating direct contact with patients’ body fluids; laying of hands by religious healers during prayers for the patient, which leads to direct body contact; sprinkling of holy water and medicine on the body of the patient. In some cases, the healers drink the medicine and hold it in their mouths from which they sprinkle to the face of the patient; and massaging the patient either while praying or applying medicine. In relation to highly infectious diseases, these processes share one thing in common – the contact with the body or body fluids which has the potential of enhancing the spread of viruses and other disease-causing pathogens.

> *What happens is after dying, we as 3 female healers wash the body clean so that we avoid spread of diseases to those who come and had no idea what they were suffering from. When the others come, we advise them to avoid touching the person because they were sick.*
>
> *Religious healer KII3, Homabay The practice of handling waste*

Interview data show that religious and traditional healers do not have the necessary knowledge and capacity to handle patients’ wastes. These wastes include vomit, blood, sputum, faecal matter, and general dirt from clothes. When asked how they dumped the wastes, most traditional healers mentioned that they dump them into a pit latrine in their compounds or bury them into the soil. Others dump or wash the wastes in water bodies that are communally used in the communities e.g., the lake, rivers, or water ponds. For example, when the traditional healer, was asked how he cleans the blankets that he uses to provide inhalation treatment to his patients, he said:

> *My wife must wash them in the lake. I also keep them in my house because it’s never anything threatening that can cause serious issues in the event the blankets are stored in the house or anything that has the potential of affecting my children*
>
> Traditional healer KII6, Homabay

This example demonstrates lack of knowledge and capacity of handling waste from patients, as pathogens causing highly infectious diseases can leak through the poor waste-handling practices into the community leading to widespread infection and overstretching management measures.

#### Healing paraphernalia

Traditional and religious healing have many paraphernalia depending on the kind of treatment given. These can range from containers for bathing, containers for drinking medicines, sharp objects for cutting or piercing the skin; blankets for inhalation; etc. Participants reported that there was widespread sharing of healing paraphernalia e.g., the same basin or bucket for bathing patients or the one calabash for drinking medicines by different patients and the same blanket for inhalation processes. Healers justified the sharing of paraphernalia in many ways namely, that it is cumbersome for patients to be bringing their own materials as they come for healing services; and that some of the paraphernalia are consecrated or spiritually dedicated for use and this is also central to the healing of a patient. Sharing of healing paraphernalia has the risks of passing infections from one patient to another.

However, other healers asked the patients to bring their own personal effects like basin, cups and plates for use during their stay as reported in the excerpt below:

> *aside, yes, he/she vomits there when it is done you tell their people now wash, they will bring their basin, if they come I tell them to buy their basin and utensils they will use to eat. He stayed three weeks and he was healed.*
>
> *Traditional healer KII1, West Pokot*

The preparation,storage and administration of the medicines demonstrate poor observation of hygiene. For example, traditional healer, described one of how he does his healing processes in the following manner:

> *Yes, once you’re seated here and these used to belong to my grandfather. I then take the hoe and detach it from the handle together with a new broom […..]. Once I have it, I then instruct my client to open his/her mouth, then the medicine moves from the broom to the hoe and lands in the mouth*
>
> Traditional healer KII6, Homabay

#### Patient handling

Traditional and religious healers are generally not trained in patient handling. They rely on their experience with various illness and patients. While their experience might be sufficient for the healthcare they give, their safety and that of their patients are not guaranteed. Most religious and traditional leaders as well as their patients mentioned that the healers usually do not use personal protective equipment (PPE) such as gloves, masks, and aprons. They handle their patients with bare hands and without protection against coughs, sneezing and spillage of body fluids such as blood. This predisposes them to highly infectious diseases in case their patients would be suffering from one. Some reasons were mentioned to be contributing to the lack of personal equipment were. First, some healers mentioned that they could not afford gloves, masks, aprons, and sterilizing agents, since they usually charge little money for their services or receive payment in kind. Another reason was the lack of knowledge that a disease could be highly infectious. Education and strategic awareness creation could help address this problem. A third reason was the need to demonstrate faith and power over ailments. This was especially true for some religious healers. The interview excerpt below helps to illustrate this point:

> *For church members to bring a sick person to you as a religious healer, this means they have a strong faith in you. You can’t tell a patient to keep distance or wear a mask so that you can pray for them because automatically they will lose hope, that strong faith they came with in you. For you to pray for a sick person you have to touch them with bare hands, and you don’t have instruments, the protective measure you have is you have to wash your hands with running water afterwards*
>
> *Religious healer KII3, Homabay*

For the religious healer, handling patients without necessary protection is meant to sustain the faith of the patient, which is an integral part of the healing process. Wearing and using protective clothing or equipment reduces the faith of the patient hence can compromise healing. However, this demonstration of spiritual faith leads to direct body contact that can be counterproductive in case the patients or the healers had EVD or other highly infectious diseases like COVID-19. It creates physical and social chains through which the diseases can spread through communities to reach pandemic levels.

The risks posed by failure to use PPE during traditional and religious healing was known to some participants. For example, some patients were aware that when a traditional or a religious healer handles them without PPE, then they can be predisposed to infections, especially of highly infectious diseases. To help illustrate this point, we consider response by a former patient of a traditional healer in the interview excerpt below:

> *Yes, it would spread fast because she was handling me without gloves, gumboots, masks, she just put on her normal clothes, she had no apron the way nurses usually have one when delivering a baby, all these would have facilitated the spread of this disease.*
>
> *Patient of a traditional healer KII3, Homabay*

Some traditional healers reported using PPEs in their healing activities. For example, a traditional birth attendant mentioned that she wears gloves when massaging pregnant women and when they accidentally come on labour and give birth at their homes she ensured that the birthing conditions was clean The interview excerpt below helps to illustrate this point:

> *You must have clean water and soap for washing hands before and after. The patient also has to be clean. The days we used to assist mothers to give birth, when cutting the umbilical cord, someone can give birth to a baby who is not very clean, so you have to wash your hands properly.*
>
> *Traditional Healer KII1, Homabay*

These relatively safe practices were reported to be as result of the collaboration between public health facilities and traditional birth attendants in the county. That the traditional birth attendant in the excerpt above intentionally ensures cleanliness and personal protection for herself and the patient is a good sign that increased awareness and education about safety during patient care can be fruitful endeavour.

## Discussion, conclusion and recommendations

Social, cultural and religious forces have historically influenced patterns of disease exposure, health-seeking behaviours, reporting of cases and the uptake of interventions. (Buckee et al., 2021). In particular, for highly infectious diseases, culturally mediated behaviors have been observed to partly shape transmission in populations which are socially organized by those cultures (Baye et al., 2021). Theoretically, cultural beliefs and practices, and health-seeking behaviours that they inspire connect with highly infectious diseases through a looping effect, leading to serious implications on outbreak prevention and response (Hacking 1995; 2007). This paper has considered traditional and religious beliefs and practices that mediate people’s response to highly infectious diseases in western Kenya and shown how looping effects emerge from those culture disease interactions.

The analysis reveals a dualistic diseases etiology, in that causes of diseases were associated with a mix of biomedical and sociocultural factors. The dual diseases etiology shapes two kinds of diseases – hospital diseases (caused by biomedical factors) and non-hospital diseases (caused by social-cultural factors). Diseases that exhibited abnormal symptoms, which are often common with highly infectious diseases e.g. hemorrhagic fevers were associated with customs and beliefs such as witchcraft, repercussions of sins and deviance from societal and cultural expectations. These observation are not new though, but had already characterized community interactions with other communicable diseases such HIV/AIDS (Odiwuor 2004; Ayikukwei et al. 2008), measles (Ouko 1998; Geissler 1998) and Schistosomiasis (Kangunyu 1997; Musuva et al. 2014) in western Kenya. It has taken great efforts of health education initiatives to change the cultural perceptions communities towards the causes of the diseases.

The dual disease etiology has affected community health-seeking behaviours, where they first seek intervention of traditional and religious healers for diseases believed to be occurring from socio-cultural causes. The practice in turn can delay appropriate management measures including diagnosis, isolation and treatment, leading to faster and wider spread of the disease and greater symptom severity for patients, especially for highly infectious diseases (see fir example Awah et al 2015, for Ebola spread in West Africa).

Cultural beliefs about how diseases is caused forms a loop with the disease in the manner in which they influence health-seeking behaviours. Non-hospital diseases were believed to be only treatable by traditional or religious healers. Interviews indicated that it was highly possible for communities to consider some highly infectious diseases as non-hospital (especially those that people are unfamiliar with). This makes the traditional and religious healers potential first line caregivers in times of outbreak of highly infectious diseases. DeVries and others (2016) have made similar conclusions from their work on community experiences with the Ebola outbreak in Uganda, where they observed that traditional healing places were amongst the first mile epidemic spaces because they were the first to try to treat patients. Prioritizing traditional and religious health-seeking in times of outbreak of highly infectious diseases will most likely delay appropriate treatment and compromise preventive response. That is, patients will only get to the formal health facilities when they are already too sick to be helped with ease and at low costs, and after they have spread the diseases to their handlers at the traditional healing places.

Preferences to traditional healers were rationalized through beliefs that the causes of those diseases are shaped by sociocultural factors and previous dissatisfaction with conventional medicine. Haque et. al. (2016), for example, suggests that the values that people derive from religious and cultural beliefs are also associated with lower trust in conventional medicine that could increase fear, which consequently lead to a more conspiratorial thinking and reduced willingness to take protective measures. For example, it was reported that some women believed that injection treatment given to children with measles at clinics prevent rashes related to the disease to appear on the body, leading children to die of a treatable disease. The findings here further explain the gaps in the knowledge of both the healer and those seeking healing in a context where measles itself is already viewed as a disease caused by sociocultural factors. This finding illuminates a need for a culturally centered health education to enhance the uptake or choice of the available interventions, especially for highly infectious diseases.

A further looping effect happens through the healing practices employed by the traditional and religious healers (Alwala, 2020). These healing practices were found to be predisposing healers, their families and their patients to increased risk of infections due to lack of protective gears and limited knowledge and capacity to handle patient’s wastes. Lack of personal protection was especially associated with inability to afford, but also the need to enhance trust and faith in the healer. Altogether, these practices increased chances of disease spreading to the healers and many patients seeking their healing services, as have been recorded in West Africa (Alexander et al., 2015; Falade, 2016; Richards et al., 2020) and Uganda (LaBrunda and Amin, 2020). Despite the challenges, popularity of these healing practices shows no decline since they are generally passed from one generation to another (Haque et al., 2016). Being the first contact people in cases of a disease outbreak, we concur with De Vries and others that interventions for highly infectious diseases like EVD should therefore not ignore, stigmatize or undermine these informal caregivers as they play a central role in the first mile strategies (De Vries et al., 2016). To achieve this goal, we contend with the argument that public health communication and education on highly infectious diseases e.g. EVD should be holistic in a manner that takes into consideration the sociocultural aspects (Manderson 1998; Sastry and Dutta 2017). We contend with Hussen et al. (2020) on the need for cultural competency in designing health communication and education programs for highly infectious diseases.

To conclude, we recast the argument that recognizing the looping effects of beliefs and how they interact with management of highly infectious diseases and need to develop a transdisciplinary approach to management of infectious disease (Parkes et., 2005). We suggest that it is advisable to meet the challenges posed by highly infectious diseases in a holistic manner because socioeconomic and cultural factors are deeply enmeshed with outbreaks of those diseases (Phua 2015). We make the following recommendations to this end:-

1. There is a need for public health education and awareness on the various highly infectious diseases e.g. hemorrhagic fevers, acute infectious respiratory diseases etc. in western Kenya and similar contexts.
2. Public health communication and education on highly infectious diseases should be culturally-centered and consider the existing disease etiologies within communities.
3. Agencies and public health professionals responding to outbreaks of highly infectious diseases should acquire cultural competencies about the contexts that they seek to respond in.
4. There is need for formulation and implementation of policies and practices to integrate alternative healing actors e.g. traditional and religious healers into the formal healthcare.

## Data Availability

All the qualititative datasets reported in the manuscript will be deposited in an appropriate public repository as indicated by PLOS data policy

## Acknowledgments

First, we convey our heartfelt thanks and gratitude to all participants who provided the data and information from which this article draws its analysis and conclusions. These participants include, religious healers, traditional healers, patients of both the healers, participants of participatory workshops (chiefs, community health promoters, public health officers and police officers from Homabay, Bungoma and West Pokot counties).

We thank the Kenya Red Cross Society staff for their outstanding contribution in ensuring successful implementation and conclusion of this assignment. Special thanks to Ms.Sharon Ayodi, Mr. Samwel Omondi, Ms. Scholastica Jelangat and Ms. Margaret Achieng for their valuable support and logistical coordination through this activity.

## Footnotes

1 See for example https://www.kenyanews.go.ke/cholera-cases-in-homa-bay-county-on-the-rise/ and https://www.standardmedia.co.ke/health/health-science/article/2001473552/cholera-outbreak-one-person-dead-73-hospitalised-in-homa-bay

## References

Adu-Gyamfi, S., & Anderson, E. (2019). Indigenous medicine and traditional healing in Africa: a systematic synthesis of the literature. *Philosophy*, Social and Human Disciplines, 1, 69–100.

Airhihenbuwa, C. O., Ford, C. L., & Iwelunmor, J. I. (2014). Why culture matters in health interventions: lessons from HIV/AIDS stigma and NCDs. Health Education & Behavior, 41(1), 78–84.

Alexander, K. A., Sanderson, C. E., Marathe, M., Lewis, B. L., Rivers, C. M., Shaman, J., & Eubank, S. (2015). What factors might have led to the emergence of Ebola in West Africa?. PLoS neglected tropical diseases, 9(6), e0003652.

Alwala, B. (2020). What has science to do with religion? A looming challenge of traditional and religious practices on curbing the spread of COVID-19 pandemic in kenya. East African Journal of Traditions, Culture and Religion, 2(1), 23–33.

Awah, P. K., Boock, A. U., & Kum, K. A. (2015). Ebola Virus Diseases in Africa: a commentary on its history, local and global context. The Pan African Medical Journal, 22(Suppl 1).

Ayikukwei, R., Ngare, D., Sidle, J., Ayuku, D., Baliddawa, J., & Greene, J. (2008). HIV/AIDS and cultural practices in western Kenya: the impact of sexual cleansing rituals on sexual behaviours. Culture, health & sexuality, 10(6), 587–599.

Balde, A. (2016). How elements of culture have contributed to the construction of health meanings in regards to the 2014 Ebola outbreak (Doctoral dissertation).

Bayeh, R., Yampolsky, M. A., & Ryder, A. G. (2021). The social lives of infectious diseases: Why culture matters to COVID-19. Frontiers in Psychology, 12, 648086.

Boahen, O., Owusu-Agyei, S., Febir, L. G., Tawiah, C., Tawiah, T., Afari, S., & Newton, S. (2013). Community perception and beliefs about blood draw for clinical research in Ghana. Transactions of The Royal Society of Tropical Medicine and Hygiene, 107(4), 261–265.

Borg, M. A. (2014). Cultural determinants of infection control behaviour: understanding drivers and implementing effective change. Journal of Hospital Infection, 86(3), 161–168.

Buckee, C., Noor, A.,and Sattenspiel, L. (2021). Thinking clearly about social aspects of infectious disease transmission. Nature, 595(7866), 205–213.

Chebii, W. K., Muthee, J. K., & Kiemo, K. (2020). The governance of traditional medicine and herbal remedies in the selected local markets of Western Kenya. Journal of ethnobiology and ethnomedicine, 16(1), 1–24.

De-Vries, D. H, Rwemisisi, J. T., Musinguzi, L. K., Benoni, T. E., Muhangi, D., Groot, M. D. & Pool, R. (2016). The first mile: community experience of outbreak control during an Ebola outbreak in Luwero district, Uganda.

Geissler, P. W. (1998). ‘Worms are our life’, part I: understandings of worms and the body among the Luo of western Kenya. Anthropology & Medicine, 5(1), 63–79.

Geissler, P. W. (2005). ‘Kachinja are coming!’: encounters around medical research work in a Kenyan village. Africa, 75(2), 173–202.

Hacking, I. (1995). The looping effects of human kinds. In D. Sperber, D. Premack, & A. J. Premack (Eds.), Causal cognition: A multidisciplinary debate (pp. 351–394). Clarendon Press/Oxford University Press.

Hacking, I. (2007). Kinds of people: Moving targets. In Proceedings-British Academy (Vol. 151, p. 285). Oxford University Press Inc..

Harrison, I. E., & Airhihenbuwa, C. O. (1993). Traditional medicine in Africa: Past, present, and future. Health and health care in developing countries: Sociological perspectives, 122–34.

Haque, M. I., Chowdhury, A. A., Shahjahan, M., & Harun, M. G. D. (2018). Traditional healing practices in rural Bangladesh: a qualitative investigation. BMC complementary and alternative medicine, 18, 1–15.

Hussen, S. A., Kuppalli, K., Castillo-Mancilla, J., Bedimo, R., Fadul, N., & Ofotokun, I. (2020). Cultural competence and humility in infectious diseases clinical practice and research. The Journal of infectious diseases, 222(Supplement_6), S535–S542.

Jones, J. (2011). Ebola, emerging: The limitations of culturalist discourses in epidemiology. The Columbia University Journal of Global Health, 1(1), 1–6.

Kagunyu, A. W. (1997). Perceptions of haematuria among the Luo of” Bondo division, Siaya district, kenya“ (Doctoral dissertation).

Kapiriri, L., & Ross, A. (2020). The politics of disease epidemics: a comparative analysis of the SARS, Zika, and Ebola outbreaks. Global Social Welfare, 7(1), 33–45.

Kinsman, J. (2012). “A time of fear”: local, national, and international responses to a large Ebola outbreak in Uganda. Globalization and health, 8, 1–12.

Kirmayer, L. J., & Sartorius, N. (2007). Cultural models and somatic syndromes. Psychosomatic medicine, 69(9), 832–840.

Kozlov, M. (2022). Ebola outbreak in Uganda: how worried are researchers?. Nature.

Kwanya, T. (2020). Stigmatisation of indigenous knowledge: The case of night-running in western Kenya. Journal of Religion in Africa, 48(4), 376–392.

LaBrunda, Michelle, and Naushad Amin. “The emerging threat of Ebola.” Global Health Security: Recognizing Vulnerabilities, Creating Opportunities (2020): 103–139.

Lucy, M., David, B., Lydia, A., Janet, K., & Patricia, K. (2022). Cultural practices resilience in the wake of COVID-19 among communities in Western Kenya.

Manderson, L. (1998). Applying medical anthropology in the control of infectious disease. Tropical Medicine & International Health, 3(12), 1020–1027.

Manson, S. M. (2020). The role of culture in effective intervention design, implementation, and research: Its universal importance. Prevention Science, 21(Suppl 1), 93–97.

Mbuthia, G. W., Olungah, C. O., & Ondicho, T. G. (2018). Health-seeking pathway and factors leading to delays in tuberculosis diagnosis in West Pokot County, Kenya: A grounded theory study. PloS one, 13(11), e0207995

McMichael, A. J. (2004). Environmental and social influences on emerging infectious diseases: past, present and future. Philosophical Transactions of the Royal Society of London. Series B: Biological Sciences, 359(1447), 1049–1058.

Musuva, R. M., Awiti, A., Omedo, M., Ogutu, M., Secor, W. E., Montgomery, S. P., … & Mwinzi, P. N. (2014). Community knowledge, attitudes and practices on schistosomiasis in western Kenya-the SCORE Project. The American journal of tropical medicine and hygiene, 90(4), 646.

Odiwuor, W. H. (2004). Indigenous Knowledge and Beliefs Among the Luo in Kenya: A Critical Inquiry into Family Life Education in the Prevention of HIV/AIDS. Inquiry: Critical Thinking Across the Disciplines, 23(4), 19–24.

Oey, E., & Rahardjo, B.S. (2021). Does culture influence our ways in handling COVID-19?. International Journal of Sociology and Social Policy, 41(11/12), 1149–1169.

Okafor, S. O., Jennifer, O. E., Nwokoma, U. B., Chuke, N. U., & Onah, S. O. (2022). Indigenous health systems and the management of infectious diseases: A study of the emergence of the COVID-19 pandemic among the Igbo of southeast Nigeria. In Indigenous Health and Well-Being in the COVID-19 Pandemic (pp. 110–129). Routledge.

Otieno, O. S., & Onyango, W. P. (2018). Influence of Socio-Culture on the Empowerment of Persons with Disabilities In Rachuonyo South-Homa-Bay County, Kenya. IOSR Journal of Humanities & Social Science, 23(8), 81–103.

Ouko, S. A. (1998). The influence of socio-culturaland economic factors on health seeking behaviour: the case of measles in children in rarieda division, siaya district’ (Doctoral dissertation, University of Nairobi).

Park, C. (2020). Traditional funeral and burial rituals and Ebola outbreaks in West Africa: a narrative review of causes and strategy interventions. Journal of Health and Social Sciences, 5(1), 073.

Parkes, M. W., Bienen, L., Breilh, J., Hsu, L. N., McDonald, M., Patz, J. A., … & Yassi, A. (2005). All hands on deck: transdisciplinary approaches to emerging infectious disease. EcoHealth, 2, 258–272.

Phua, K. L. (2015). Meeting the challenge of Ebola virus disease in a holistic manner by taking into account socioeconomic and cultural factors: the experience of West Africa. Infectious Diseases: Research and Treatment, 8, IDRT-S31568.

Richards, P., Mokuwa, G. A., Vandi, A., Mayhew, S. H., & Ebola Gbalo Research Team. (2020). Re-analysing Ebola spread in Sierra Leone: the importance of local social dynamics. PloS one, 15(11), e0234823

Rodriguez-Dod, E. C., Marty, A. M., & Marty-Nelson, E. M. (2016). Tears in heaven: Religiously and culturally sensitive laws for preventing the next pandemic. Cath. UL Rev., 66, 117.

Ryder, A. G., & Chentsova-Dutton, Y. E. (2015). 16 Cultural-Clinical Psychology. *Re-Visioning Psychiatry: Cultural Phenomenology*, Critical Neuroscience, and Global Mental Health, 400.

Sastry, S., & Dutta, M. J. (2017). Health communication in the time of Ebola: A culture-centered interrogation. Journal of health communication, 22(sup1), 10–14.

Schaller, M., & Murray, D. R. (2010). Infectious diseases and the evolution of cross-cultural differences. Evolution, culture, and the human mind, 243–56.

Schiavo, R. (2013). Health communication: From theory to practice (Vol. 217). John Wiley & Sons.

Shultz, J. M., Cooper, J. L., Baingana, F., Oquendo, M. A., Espinel, Z., Althouse, B. M., . & Rechkemmer, A. (2016). The role of fear-related behaviors in the 2013–2016 West Africa Ebola virus disease outbreak. Current psychiatry reports, 18, 1–14.

Victor, G. S., & Ahmed, S. (2019). The importance of culture in managing mental health response to pandemics. Psychiatry of pandemics: A mental health response to infection outbreak, 55-64.

Wonnah, S. Z. (2018). Myths, Risks, and Ignorance: Western Media and Health Experts’ Representations of Cultures in Ebola-Affected West African Communities (Doctoral dissertation, East Tennessee State University).

World Health Organization. (2013). *WHO traditional medicine strategy: 2014-*2023. World Health Organization.

